# Published Research on the Human Health Implications of Climate Change from 2012-2019: a Cross-Sectional Study

**DOI:** 10.1101/2022.10.01.22280596

**Authors:** Victoria L. Bartlett, Ravi Gupta, Joshua Wallach, Kate Nyhan, Kai Chen, Joseph S. Ross

**Affiliations:** Yale School of Medicine, New Haven, CT, USA; Johns Hopkins University School of Medicine, Baltimore, MD; Department of Environmental Health Sciences, Yale School of Public Health, New Haven, CT; Collaboration for Research Integrity and Transparency (CRIT), Yale Law School, New Haven, CT; Harvey Cushing/John Hay Whitney Medical Library, Yale University, 333 Cedar Street, New Haven, CT; Section of General Internal Medicine and National Clinician Scholars Program, Yale School of Medicine, New Haven, CT; Department of Health Policy and Management, Yale University School of Public Health, New Haven, CT; Center for Outcomes Research and Evaluation, Yale-New Haven Hospital, New Haven, CT

## Abstract

**Importance:** Climate change is one of the most pressing global public health issues and is increasingly gaining attention from governments and researchers as a critical policy and research focus. While research on the effects of climate change on human health has grown significantly over the past few decades, there remain known gaps in research on non-physical health topics, like mental health, and in research on low-income countries.

**Objective:** To better understand the current state of research on the human health effects of climate change, including exposures, health conditions, populations, areas of the world studied, funding sources, and publication characteristics, with a focus on topics that are relevant for vulnerable populations.

**Evidence Review:** We searched the National Institute of Environmental Health Sciences (NIEHS) Climate Change and Human Health Literature Portal, a curated bibliographic database of global peer-reviewed research and gray literature on the science of climate impact on human health, to identify peer-reviewed original research investigating the health effects of climate change published from 2012 to 2019. The database combines searches of multiple search engines including PubMed, Web of Science, and Google Scholar and includes added-value expert tagging of climate change exposures and health impacts. We filtered our search by year published, limited to original research articles published in English. After identifying all original research articles, we selected a 5% random sample to manually perform a detailed characterization of research topics and publication information.

**Findings:** There were 7082 original research articles published between 2012 and 2019, and the number of articles increased by 23% annually. In our random sample of 348 articles, we found that there were several gaps in research topics that are particularly relevant to vulnerable populations, such as those in the Global South (159; 45.7%) and the elderly (55; 15.8%). Additionally, fewer first authors were from the Global South (110; 31.6%), which may in part explain why there is disproportionally less research focusing on these countries.

**Conclusions and Relevance:** Our results help elucidate gaps in research that, once addressed, may help us better understand and mitigate some of the most devastating effects of climate change on human health.

## Introduction

Climate change is one of the most pressing public health threats of the 21^st^ century, contributing to over 250,000 deaths each year.^1,2^ Global temperatures were 1.09 degrees Celsius warmer in 2011-2020 compared to 1850-1900, changing oceanic and atmospheric systems and contributing to rising sea levels, wild fires, flooding, droughts, and other extreme weather events.^3^ The environmental effects of climate change have increasingly important and varied implications for human health through multiple pathways. Climate change has direct and indirect effects on morbidity and mortality, food and water insecurity, and social and economic challenges, such as financial stability and human displacement.^3,4^ The type and severity of climate change effects vary by region, and countries in the Global South that historically least contributed to anthropogenic climate change are often disproportionately affected.^1,4-7^ Climate-related stressors exacerbate existing inequalities within regions and disproportionately affect vulnerable and oppressed populations, including those living in poverty.^8,9^ Understanding the local economic, social, and environmental factors that uniquely influence a population’s vulnerability to climate change is critical in order to effectively and equitably address this threat across the world.^10^

Governments and researchers have been paying increasing attention to understanding the human health implications of climate change.^11^ In 2015, the same year that the Paris Agreement was adopted by 196 countries pledging to limit global warming to below 1.5 degrees Celsius compared to pre-industrial levels, The Lancet created a Commission on Health and Climate Change that publishes a yearly report on the topic.^1,12,13^ Beyond the Lancet Commission, articles on the human health effects of climate change increased eight-fold between 2007 and 2019.^1^ According to a recent study that used machine learning to systematically map the existing literature on climate change and human health, the most commonly studied topics were heat-related effects on health.^14^ There were major gaps in research on non-physical health topics like mental health, and the research was disproportionately produced by high-income countries, consistent with prior studies.^1,3,6,15^

Prior studies have not focused on characterizing the topics, publication characteristics, and funding sources of existing literature on the effects of climate change on human health that are relevant to countries in the Global South. We conducted a cross-sectional study of a representative sample of original research studies on the human health effects of climate change published between 2012 and 2019 to better understand the current state of research examining the effects of climate change on health, including manual review of exposures, health conditions, populations, geographies studied, funding sources, and publishing characteristics, with a focus on differentiating what has been studied in the Global South and in the Global North. Details of the existing literature on climate change and human health and the topical emphases of prior studies can help to direct future research and policy efforts to topics and geographies with the greatest need.

## Methods

### Search Strategy

We searched the National Institute of Environmental Health Sciences (NIEHS) Climate Change and Human Health Literature Portal, a curated bibliographic database of global peer-reviewed research and gray literature on the science of climate impact on human health, to identify peer-reviewed original research investigating the health effects of climate change published from 2012 to 2019.^16,17^ The database combines searches of multiple search engines including PubMed, Web of Science, and Google Scholar and includes added-value expert tagging of climate change exposures and health impacts. This approach of identifying studies via screening records from an evidence hub, rather than relying on keyword searches in traditional bibliographical databases, aligns with our goal of characterizing a random sample of literature recognized as addressing climate change and human health.

We filtered our search by year published, limited to original research articles (defined as an article in a peer-reviewed journal containing original data) published in English. Search results were downloaded on June 20, 2021. After identifying all original research articles, we selected a 5% random sample to manually perform a detailed characterization of research topics and publication information. One investigator (VLB) read the title and abstract to verify that the articles examined associations between climate change and health. One investigator (VLB) read the abstracts, and full text, if necessary, of each article in the sample in order to characterize the below study characteristics; any uncertainties were clarified with another investigator (JSR).

### Study Characteristics

For each article, we abstracted the climate change exposure(s), health effect(s), geographic location(s), and population(s) studied. We also determined the publication year, journal impact factor, number of authors, location of the first author’s affiliation, and funding source(s). Climate change exposure(s) were categorized into 8 groups: general exposure; air pollution; extreme weather; food and water quality and security; human conflict/displacement; indoor environment; seasonality, temperature, and meteorological factors; and other (**Table 1**). Health effect(s) were categorized into 19 groups (**Table 1**). These groups were based on the categories in the NIEHS portal. We validated the portal’s tagging of climate change exposures and health effects and altered it if we disagreed. Articles were categorized as studying general health effects if they did not name a specific health effect but health was mentioned generally and the climate change exposure studied was one that clearly affects health (e.g., water quality, food security, heat waves). Geographic location(s) was categorized as North America, South America, Europe, Asia, Africa, Australasia, Antarctica, global, or unspecified. Articles categorized as global were those in which specific geographies were not stated and/or global data were analyzed in aggregate. We also characterized whether the article studied at least one country in the Global South, according to the United Nations’ definition of the Global South.^18^ We counted an article in the Global South category even if it had a global focus, as long as it was relevant to at least one Global South country.

**Table 1:** Research topics of original research articles studying the association between climate change and health, 2012 to 2019.

|  | **Number of articles (% of sample<sup>a</sup>)** | **Annual increase (%)<sup>b</sup>** | **Articles with Global South focus (%)<sup>c</sup>** |
| --- | --- | --- | --- |
| **Total** | 348 (100.0) | 26 | 159 (45.7) |
| **Climate change exposures<sup>d</sup>** |  |  |  |
| General exposure | 67 (19.3) | 13 | 34 (50.7) |
| Air pollution | 41 (11.8) | 41 | 14 (34.1) |
| Extreme weather | 113 (32.5) | 29 | 40 (35.4) |
| Food & water quality & security | 53 (15.2) | 22 | 32 (60.4) |
| Human conflict/displacement | 8 (2.3) |  | 8 (100) |
| Indoor environment | 7 (2.0) |  | 0 (0.0) |
| Seasonality, temperature, & meteorological factors | 228 (65.5) | 34 | 101 (44.3) |
| Other | 20 (5.7) |  | 9 (45.0) |
| **Health effects** |  |  |  |
| Cancer | 3 (0.9) |  | 2 (66.7) |
| Cardiovascular | 19 (5.5) |  | 9 (47.4) |
| Dermatological | 1 (0.3) |  | 0 (0.0) |
| Developmental | 3 (0.9) |  | 1 (33.3) |
| Diabetes/obesity/overweight | 3 (0.9) |  | 0 (0.0) |
| Economic | 33 (9.5) | -8 | 21 (63.6) |
| General Health | 65 (18.7) | 12 | 36 (55.4) |
| Infectious disease | 89 (25.6) | 19 | 54 (60.7) |
| Injury | 5 (1.4) |  | 2 (40.0) |
| Malnutrition | 7 (2.0) |  | 6 (85.7) |
| Medical visit | 11 (3.2) |  | 3 (27.3) |
| Mental health and well-being | 9 (2.6) |  | 3 (33.3) |
| Morbidity and mortality | 49 (14.1) | 32 | 13 (26.5) |
| Neurological | 5 (1.4) |  | 2 (40.0) |
| Reproductive | 7 (2.0) |  | 2 (28.6) |
| Respiratory | 22 (6.3) | 20 | 5 (22.7) |
| Temperature-related health | 42 (12.1) | 37 | 4 (9.5) |
| Urologic | 7 (2.0) |  | 2 (28.6) |
| Other health | 28 (8.0) | 7 | 13 (46.4) |
| **Geographic area** |  |  |  |
| Unspecified | 8 (2.3) |  | NA |
| Global | 10 (2.9) |  | 10 (100.0) |
| North America | 74 (21.3) | 22 | 8 (10.8) |
| South America | 19 (5.5) |  | 19 (100.0) |
| Europe | 79 (22.7) | 17 | 5 (6.3) |
| Asia | 110 (31.6) | 69 | 97 (88.2) |
| Africa | 31 (8.9) | 29 | 31 (100.0) |
| Oceania | 32 (9.2) | 14 | 4 (12.5) |
| **Region** |  |  |  |
| Global South<sup>e</sup> | 159 (45.7) | 45 | NA |
| Global North | 181 (52.0) | 20 | NA |
| **Population focus** |  |  |  |
| Elderly | 55 (15.8) | 22 | 16 (29.1) |
| Adults | 56 (16.1) | 31 | 26 (46.4) |
| Children | 98 (28.2) | 31 | 38 (38.8) |
| Not specified | 222 (63.8) | 24 | 108 (48.6) |

Abbreviations: NA, Not Applicable
<sup>a</sup> Sample is 5% of total number of articles. <sup>b</sup> Annual increase not calculated if total less than or equal to 20.
<sup>c</sup> Denominator was number of articles in the sample. <sup>d</sup> Climate change exposures: extreme weather-related event or disaster (i.e.,
4 extreme precipitation events, tropical cyclones, floods, droughts, wildfires, heat waves, cold waves); food and water quality and
- 1 security; human conflict and displacement; indoor environment; air pollution; seasonality, temperature, and meteorological factors; - 2 general exposures; other (including ecosystem change, solar radiation, sea level rise, and sea surface oscillation). <sup>e</sup>According to the United Nations' Finance Center for South-South Cooperation.

Population focus(es) were categorized as children, adults, elderly, or not specified. Funding source(s) were categorized as government, non-profit, for-profit, academic, and not specified. The journal’s 2020 impact factor was obtained from Web of Science Journal Citation Reports, and categorized as 0-4.99, 5.00-9.99, or 10.00 and greater.

### Statistical Analyses

We used descriptive statistics to characterize the 5% sample. For categorical variables, we reported the percentage of articles within each category. For continuous variables, we reported the median and interquartile range. We also used the same methodology to characterize the subset of articles studying the Global South. Within the random sample, we calculated the average annual percentage change in the number of articles for each variable if the total was greater than 20 articles. Data were analyzed in Excel, version 16.54 (Microsoft).

## Results

### Number of articles and years published

Based on search results from the NIEHS portal on July 20, 2021, we identified 7082 original research articles published between 2012 and 2019: 297 (4.2%) were published in 2012, 187 (2.6%) in 2013, 845 (11.9%) in 2014, 831 (11.7%) in 2015, 1103 (15.6%) in 2016, 1196 (16.9%) in 2017, 1376 (19.4%) in 2018, and 1247 (17.6%) in 2019, representing a 23% annual growth (**Figure 1**).

**Figure 1:**
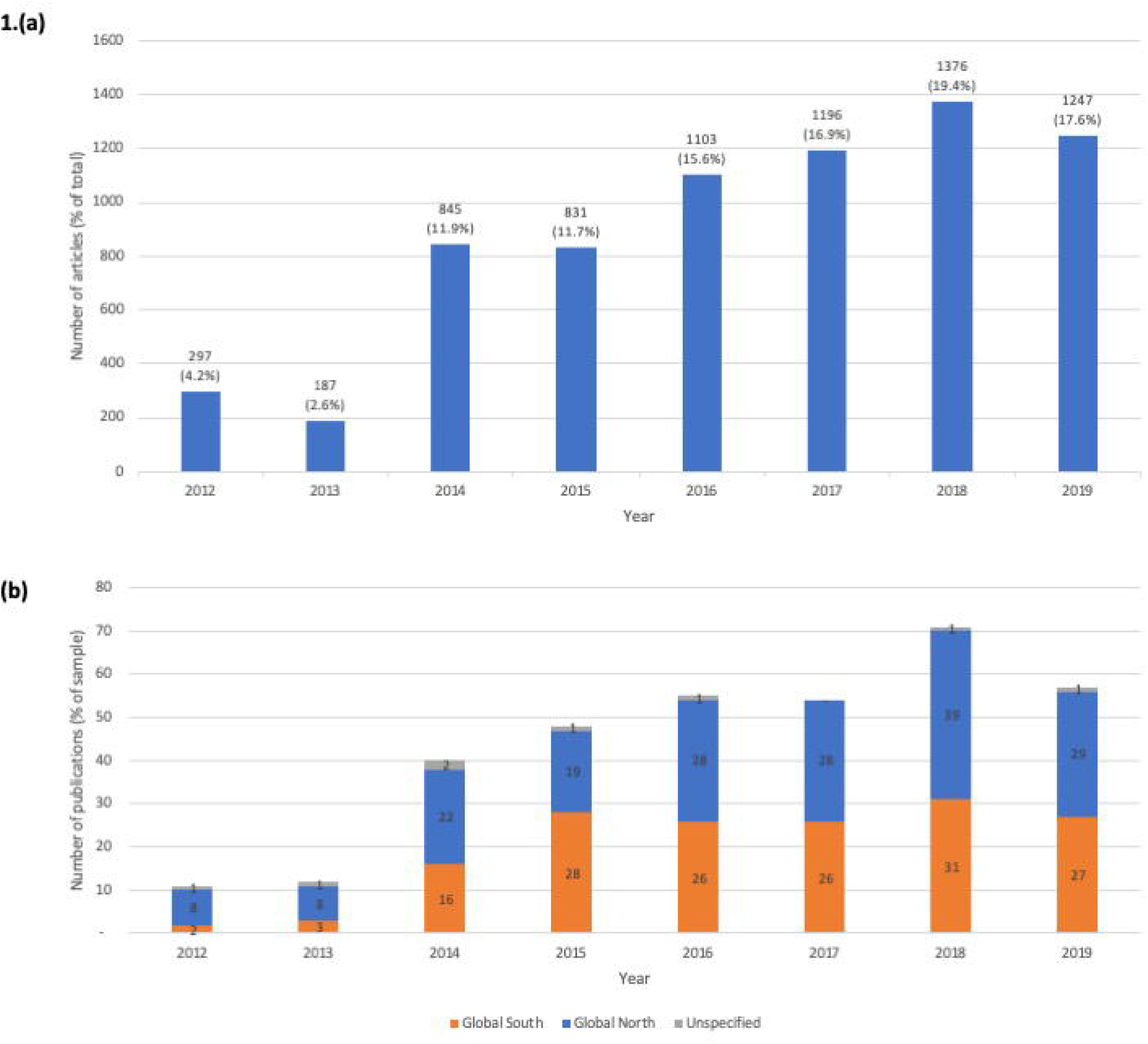
Original research articles studying the association between climate change and health, 2012-2019, **(a)** in total; **(b)** in 5% sample

In the random 5% sample of 354 articles, 6 (1.7%) were excluded because they were not directly related to health and/or climate change. Of the remaining 348 articles, 11 (3.2%) were published in 2012, 12 (3.4%) in 2013, 40 (11.5%) in 2014, 48 (13.8%) in 2015, 55 (15.9%) in 2016, 54 (15.8%) in 2017, 71 (20.4%) in 2018, and 57 (16.4%) in 2019. This represented a 26% average annual growth in total number of articles published.

### Climate change exposures studied

Among the 348 articles, 228 (65.5%) studied seasonality, temperature, and meteorologic factors, 101 (44.3%) of which studied the Global South; 113 (32.5%) studied extreme weather, 40 (35.4%) of which studied the Global South; and 53 (15.2%) studied food and water quality and security, 32 (60.4%) of which studied the Global South (**Table 1**). The number of articles studying air pollution grew by 41% annually, while those studying seasonality, temperature & meteorologic factors and extreme weather grew by 34% and 29% annually, respectively (**Figure 2a**).

**Figure 2:**
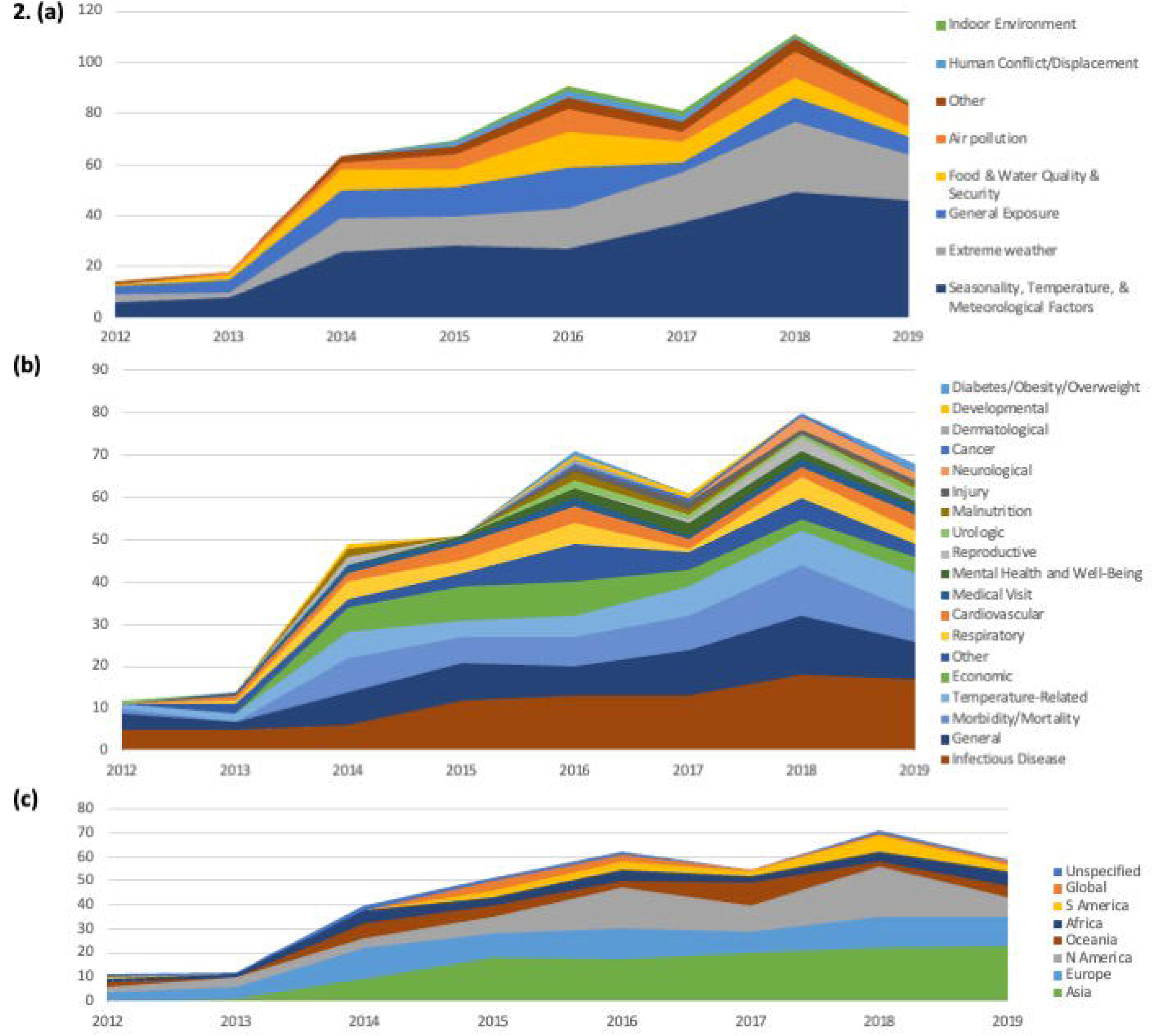
**(a)** Climate change exposure(s); **(b)** health effect(s); and **(c)** geographies studied in original research articles studying the association between climate change and health, 2012 to 2019

### Health effects studied

Sixty-five (18.7%) articles studied general health, of which 36 (55.4%) studied the Global South; 89 (25.6%) studied infectious disease, of which 54 (60.7%) studied the Global South; 49 (14.1%) studied morbidity and mortality, of which 13 (26.5%) studied the Global South; 33 (9.5%) studied economic effects, of which 21 (63.6%) studied the Global South; and 9 (2.6%) studied mental health effects, of which 3 (33.3%) studied the Global South (**Table 1**). Articles studying temperature-related health effects, morbidity and mortality, respiratory effects grew by 37%, 32%, and 20% annually respectively (**Figure 2b**).

### Population geographic location studied

Asia was the most commonly studied continent (110; 31.6%), followed by Europe (79; 22.7%), North America (74; 21.3 %), Oceania (32; 9.2%), Africa (31; 8.9%), then South America (19; 5.5%). Ten (2.9%) had a global focus, and 8 (2.3%) did not specify a geography (**Table 1**). The number of articles studying Asia grew by 69% annually, those studying Africa grew by 29% annually, and those studying North America grew by 22% annually (**Figure 2c**). In 2012, only 2 of the 11 (18.2%) articles studied the Global South, whereas in 2019, 27 of 56 (48.2%) did, representing a 45% year-over-year growth (**Figure 1**). Over half of the articles did not study any country in the Global South (181; 52.0%).

### Population demographics studied

Most articles did not specify an age-related population or report the age distributions (e.g., mean or median) (222; 63.8%), while 98 (28.2%) studied adult, 56 (16.1%) pediatric, and 55 (15.8%) elderly populations (**Table 1**). Among articles with age-related information, 38 (38.8%), 26 (46.4%), and 16 (29.1%) studied adult, pediatric, and elderly populations, respectively, in the Global South.

### Author characteristics (location, funding)

The median number of authors per article was 5 (IQR, 3 – 7). The most common first first author primary affiliation location was Asia (103; 29.6%), followed by Europe (97; 27.9%), North America (93; 26.7%), Oceania (31; 8.9%), South America (14; 4.0%), and Africa (10; 2.9%). The number of articles with first author primary affiliation in Asia grew by 37% annually, in North America by 22% annually, and in Oceania by 20% annually (**Table 2**). First authors with a primary affiliation in a Global South country made up 31.6% (110) of articles, and those in a Global North country 68.4% (238) of articles. The number of articles with a first author primary afilliation in a Global South country grew by 52% annually. Of the 159 articles studying the Global South, 107 (67.3%) had first author with primary affiliation in the Global South. Of the 238 articles with first author primary affiliation in the Global North, 179 (75.2%) studied only countries in the Global North, while 52 (21.8%) studied at least one country in the Global South. The majority of authors with primary affiliation in the Global South studied countries in the Global South (107; 97.3%), with the number of such articles growing by 52% annually.

**Table 2:** Study characteristics of original research articles studying the association between climate change and health, 2012 to 2019.

|  | **Number of**<br>**articles (% of**<br>**sample<sup>a</sup>)** | **Annual**<br>**increase<sup>b</sup>**<br>**(%)** | **Articles with**<br>**Global South**<br>**focus (%)<sup>c</sup>** |
| --- | --- | --- | --- |
| **Total** | 348 (100.0) |  | 159 (45.7) |
| **Journal impact factor** |  |  |  |
| 0.00-4.99 | 11 (3.2) | - | 3 (27.3) |
| 5.00-9.99 | 83 (23.9) | 31 | 30 (36.1) |
| ≥10.00+ | 253 (72.7) | 30 | 125 (49.4) |
| **Median number of authors**<br>**(interquartile range)** | 5 (3 – 7) | - | - |
| **First author location (continent)** |  |  |  |
| North America | 93 (26.7) | 22 | 22 (23.7) |
| South America | 14 (4.0) | - | 14 (100.0) |
| Europe | 97 (27.9) | 16 | 21 (21.6) |
| Asia | 103 (29.6) | 37 | 85 (82.5) |
| Africa | 10 (2.9) | - | 10 (100.0) |
| Oceania | 31 (8.9) | 20 | 7 (22.6) |
| **First author location (region)** |  |  |  |
| Global South<sup>d</sup> | 110 (31.6) | 52 | 107 (97.3) |
| Global North | 238 (68.4) | 21 | 52 (21.8) |
| **Funding source** |  |  |  |
| Government | 223 (64.1) | 25 | 90 (40.4) |
| Non-profit | 31 (8.9) | 31 | 14 (45.2) |
| For-profit | 3 (0.9) | - | 2 (66.7) |
| Academic | 75 (21.6) | 18 | 34 (45.3) |
| Not specified | 64 (18.4) | 6 | 36 (56.3) |
| None | 26 (7.5) | 0 | 10 (38.5) |

| **Conflict of Interest** |  |  |  |
| --- | --- | --- | --- |
| Oil & gas funding | 0 (0.0) | - | 0 (0.0) |
| Pharmaceutical funding | 1 (0.3) | - | 0 (0.0) |
| No funding | 185 (53.2) | 36 | 79 (42.7) |
| Not specified | 156 (44.8) | 10 | 77 (49.4) |

<sup>a</sup>Sample is 5% of total number of articles. <sup>b</sup>Annual increase calculated if total greater than 20. <sup>c</sup> Denominator was number of articles in the sample. <sup>d</sup> According to the United Nations' Finance Center for South-South Cooperation.

The majority of articles were funded by government sources (223; 64.1%), while 75 (21.6%) were funded by academic sources, 31 (8.9%) by non-profit sources, and 3 (0.9%) by for-profit sources. Twenty-six (7.5%) articles stated they did not receive funding, while 64 (18.4%) did not specify whether or not they received funding for the work presented (**Table 2**).

### Journal characteristics

Two-hundred and fifty-three (72.7%) articles were published in journals with impact factor less than 5, of which 125 (49.4%) studied a country in the Global South. Among the 83 (23.9%) articles that were published in journals with impact factor 5 to 9.99, 30 (36.1%) studied a country in the Global South. Of the 11 (3.2%) articles published in journals with an impact factor 10 and above, 3 (27.3%) studied a country in the Global South (**Table 2**). Articles with impact factor 5 to 9.99 grew by 31% annually, and those with impact factor less than 5 grew by 30% annually.

## Discussion

In this cross-sectional evaluation of a random sample of original research articles studying the human health effects of climate change published between 2012 and 2019, we found that the number of articles published increased by 23% annually. This likely reflects the growing global research interest in the effects of climate change on human health and wellbeing, and of the crucial role of human activity in affecting climate change. Despite this growth, we found several gaps in research topics that are relevant to more vulnerable populations, such as those in the Global South and the elderly. Fewer first authors were from the Global South, which may in part explain the disproportionally less research focusing on these countries, and may be linked to research funding sources and allocation.

While articles studying the Global South have grown by 45% annually, they still constitute a minority of original research articles studying climate change and human health. However, by 2100, 77% of the world’s population is predicted to live in the Global South, where countries bear a disproportionate share of the effects of global warming.^19,20^ In addition, the distribution of climate change exposures studied reflects a bias towards those that affect higher-income countries. The two most studied topics – extreme weather and seasonality, temperature and meteorology – had disproportionately fewer articles studying the Global South. In contrast, articles studying human conflict and migration, all of which studied the Global South, made up a small proportion of all articles. The impact of climate change on human conflict and migration is especially relevant for countries in the Global South, and given that the number of climate refugees from Latin America, sub-Saharan Africa, and Southeast Asia alone is predicted to increase by 143 million people before 2050, it is critical to continue conducting research on this consequential climate change exposure and its potentially global effects.^21^

The distribution of illnesses studied also suggests disproportionate gaps in research most relevant to Global South countries. The impact of climate change on infectious diseases and general health were the most commonly studied, while mental health and economic effects were studied by fewer articles. Evidence suggests that low-income countries in the Global South in particular are the most affected by heat-related reductions in labor capacity, with 2015 average estimated losses equivalent to 3.9% to 5.9% of gross domestic product in countries such as Indonesia, India, and Cambodia.^1^ Moreover, financial stress is the strongest predictor of mental health issues after natural disasters, and people living in the Global South are at increased risk of exposure to natural disasters and poverty.^22^

Few articles studied the elderly, an especially vulnerable population, and only a quarter of these studied elderly populations in the Global South. Older adults are particularly susceptible to the effects of climate change, including extreme weather events, air quality, and infectious disease.^23^ Moreover, low-income older adults are less likely to have the resources to adapt to the effects of climate change. More research is needed to understand the unique needs of these vulnerable populations and to find geographically appropriate solutions.

Most articles’ first authors were from Global North countries, and only one fifth of them studied Global South countries. Additionally, articles studying Global South countries were disproportionately published in journals with lower impact factor. Most articles received government funding, which may be less likely to fund studies that do not focus on their own geographic region. These findings may partially explain the disproportionate gap in quality and quantity of articles studying the Global South observed. Moreover, health-related research bodies globally underfund and underemphasize research on climate change and health.^10^ There is a need for greater funding of climate change and health research, and particularly of researchers from countries in the Global South and of studies that are relevant to the people that live in these locations, which may require increased funding from academic, non-profit, and for-profit sources in the Global North.

Increasing the number and variety of research studies on the health effects of climate change may contribute to increased popular support for government engagement with climate change. Currently, only a small percentage of research on climate change focuses on its health effects.^1^ In the annual United Nations General Assembly, engagement in the health effects of climate change is high in governments of island nations who face greater risks, particularly in the Western Pacific region, while engagement is low among countries with the highest carbon dioxide emissions, including the United States and those in the European Union.^1^ Increasing research on the effects of climate change, especially on topics relevant to the Global South, may draw attention to and allow for increased investment in mitigating the health effects of climate change globally.

## Limitations

There are several limitations to this study. First, we manually characterized a random subset of all research articles. Although we did not consider all articles, the random sample should be representative of the total population of research on climate change and human health. Second, only one author screened and abstracted data, although uncertainties were discussed with a second author. Third, we relied on only one data source, though the NIEHS portal itself combines results from multiple search engines. Fourth, when characterizing the geographic location of the authors, we only considered the first author, so this may not reflect the full scope of affiliations of all co-authors. Fifth, we did not include articles published before 2012, or those that were not published in English. However, given the recent growth in this topic, the majority of research was published during the studied time period.

## Conclusion

In this characterization of original research on the health effects of climate change, we found that although the body of research has grown substantially over the last decade, there continues to be a disproportionate focus on countries in the Global North and less vulnerable populations. By increasing funding for research on neglected topics, such as human conflict and displacement, economic effects, elderly populations, and countries in the Global South, we may be better equipped to address some of the most devastating potential effects of climate change. While research in these areas is growing, it may require increased funding from entities that are not tied to certain governments or geographies, such as non-profit and academic organizations, to achieve the volume and quality of research needed to address these issues.

## Data Availability

All data produced in the present study are contained in the manuscript.

## Acknowledgements

The authors would like to acknowledge the National Institute of Environmental Health Sciences’ Climate Change and Human Health Portal’s curation of the research articles used in this study.

